# CHRONOSIG: Digital Triage for Secondary Mental Healthcare using Natural Language Processing - rationale and protocol

**DOI:** 10.1101/2021.11.23.21266750

**Authors:** Dan W Joyce, Andrey Kormilitzin, Julia Hamer-Hunt, Anthony James, Alejo Nevado-Holgado, Andrea Cipriani

## Abstract

**Background:** Accessing specialist secondary mental health care in the NHS in England requires a referral, usually from primary or acute care. Community mental health teams triage these referrals deciding on the most appropriate team to meet patients’ needs. Referrals require resource-intensive review by clinicians and often, collation and review of the patient’s history with services captured in their electronic health records (EHR). Triage processes are, however, opaque and often result in patients not receiving appropriate and timely access to care that is a particular concern for some minority and under-represented groups. Our project, funded by the National Institute of Health Research (NIHR) will develop a clinical decision support tool (CDST) to deliver accurate, explainable and justified triage recommendations to assist clinicians and expedite access to secondary mental health care.

**Methods:** Our proposed CDST will be trained on narrative free-text data combining referral documentation and historical EHR records for patients in the UK-CRIS database. This high-volume data set will enable training of end-to-end neural network natural language processing (NLP) to extract ‘signatures’ of patients who were (historically) triaged to different treatment teams. The resulting algorithm will be externally validated using data from different NHS trusts (Nottinghamshire Healthcare, Southern Health, West London and Oxford Health). We will use an explicit algorithmic fairness framework to mitigate risk of unintended harm evident in some artificial intelligence (AI) healthcare applications. Consequently, the performance of the CDST will be explicitly evaluated in simulated triage team scenarios where the tool *augments* clinician’s decision making, in contrast to traditional “human versus AI” performance metrics.

**Discussion:** The proposed CDST represents an important test-case for AI applied to real-world process improvement in mental health. The project leverages recent advances in NLP while emphasizing the risks and benefits for patients of AI-augmented clinical decision making. The project’s ambition is to deliver a CDST that is scalable and can be deployed to any mental health trust in England to assist with digital triage.

## BACKGROUND

In the United Kingdom, mental healthcare is stratified into primary (led by General Practice), secondary (community and hospital NHS Trusts) and tertiary services (e.g. secure forensic services). In 2019-2020, over 2.8 million people in England were in contact with secondary mental health (SMH) services with around 104,500 being admitted to hospital and the majority (96%, or 2.69 million) receiving care from community-based teams (NHS Digital, 2020).

Patients requiring secondary care are referred to a triage and assessment function, contained in community mental health teams (CMHTs) operating in NHS Trusts (collections of community-based clinics and inpatient hospitals). Referrals for triage and assessment are via documents written by the referring professional who is most often a general practitioner (GP), a healthcare professional in an emergency department or member of a social care organisation.

In March 2021, across England’s mental healthcare NHS Trusts, there were 404,552 new referrals (NHS Digital, 2021). Each referral requires the CMHT to instruct one of their clinicians to review the referral documentation alongside an examination of historical medical notes (e.g. if the patient is already known to SMH services) usually concluding with a discussion and decision in a multi-disciplinary team (MDT) meeting. The outcome of these processes is to reject the referral (e.g. if clinically inappropriate or the incorrect locality/service), obtain further information (e.g. from the referrer, patient and carers) or allocate to a specialty or locality team for further assessment and treatment. The cost of initial assessments arising from new referrals to SMH services was £326 million in 2018-2019 (NHS England, 2020).

Referral and triage processes lack transparency (to patients and referrers), are capricious (with CMHTs using referral criteria and thresholds inconsistently) and result in frustration from ‘referral bouncing’ (Chew-Graham et al., 2007). Attempts at improving this interface -- for example, with GPs and CMHTs using a standardised referral tool to ‘grade’ patient’s severity, risk and unmet needs -- have yielded poor results (Slade et al., 2008). Patient care is delayed because they are being triaged multiple times, by different teams, who may disagree on the most appropriate treatment team for the patient’s needs.

The delay in accessing treatment (after referral) has been labelled the “hidden waiting list” by The Royal College of Psychiatrists who identified that two-fifths of patients end up accessing emergency or crisis care (RCPsych, 2020). Displacing patients with unmet mental health needs impacts on other NHS services -- for example, in 2013-2014, there were 6.2 million emergency department attendances across England for mental health needs with around half being discharged back to community care (Baracaia et al., 2020). It is certainly true that patients being denied timely and appropriate care is largely attributable to inadequate national resourcing for mental health services (Docherty & Thornicroft, 2015) -- however, the referral and triage process is one target for process improvement where both patients and clinicians stand to benefit. To this end, this project directly addresses the strategic priorities described in the Topol review (Foley & Woollard, 2019) and the NHS Long Term Plan (NHS England, 2019) that emphasise the use of EHRs to improve and personalise care for individuals.

CHRONOSIG, which stands for CHRONOlogical SIGnature, is a National Institute of Health Research-funded project designed to improve triage by leveraging contemporary machine learning (ML) technology on information contained in large, observational electronic health record (EHR) data. Each patient’s historical EHR data captures one or more instances (or episodes) of care under mental health services. CHRONOSIG will use natural language processing (NLP) techniques to represent a patient’s longitudinal signature (or ‘fingerprint’) -- capturing their history, signs/symptoms and presenting difficulties -- and then learn associations between these signatures and triage decisions (captured as explicit structured data in patient’s EHR data). The aim is to deliver a triage clinical decision support tool (CDST) that takes as input a patient’s referral documentation, utilises existing medical notes (when available) and delivers a suggested triage outcome to assist MDTs.

## METHODS

### Overview

Figure 1 shows an outline user-interface for the clinician-facing CDST. A clinician provides a referral document and (if available) the patient’s existing EHR data (presented to the CDST as a time-ordered sequence of free-text clinical notes). The CDST computes an embedded representation of chronological free-text data (described later). This transforms the patient’s raw free-text data into a space of signatures that exposes patterns of similarity/dis-similarity to the signatures of all patients the CDST encountered during development or ‘training’ of the algorithms (Figure 1, Panel C). A follow-on classifier -- that has been trained to associate signatures with known triage outcomes -- delivers a conditional distribution of continuous triage outcome scores; for example, in Figure 1 (Panel A), scores are represented as a conditional probability distribution of triage outcomes [0.1, 0.3, 0.5, 0.1] for teams W,X,Y and Z respectively. At this stage, a recommendation rule must be applied to these scores. Figure 1 (Panel A) shows one such rule where the maximum value is taken as the *recommended* triage outcome (i.e. Team Y with score 0.5) and the *confidence* is shown as inversely proportional to the entropy (Cover, 1999) of this distribution (approximately, if the distribution is heavily ‘peaked’ around a single team, the entropy is low and the confidence high). Note that triage scores need not be mutually exclusive -- in the example shown in Figure 1, Team X might also provide benefit to the patient because the patient’s signature displayed features (for example, co-morbidity) similar to other patients treated by Team X. To assist clinicians, they can then explore features in the patient’s raw data that were *salient* and driving the classifier’s outputs (Figure 1, Panel B). These components are the foundation for a principled **AI-augmented** clinical decision making process - rather than the CDST delivering an automated triage decision, the final “decision rule” is a combination of the output of the CDST with the MDT’s clinical expertise.

**Figure 1:**
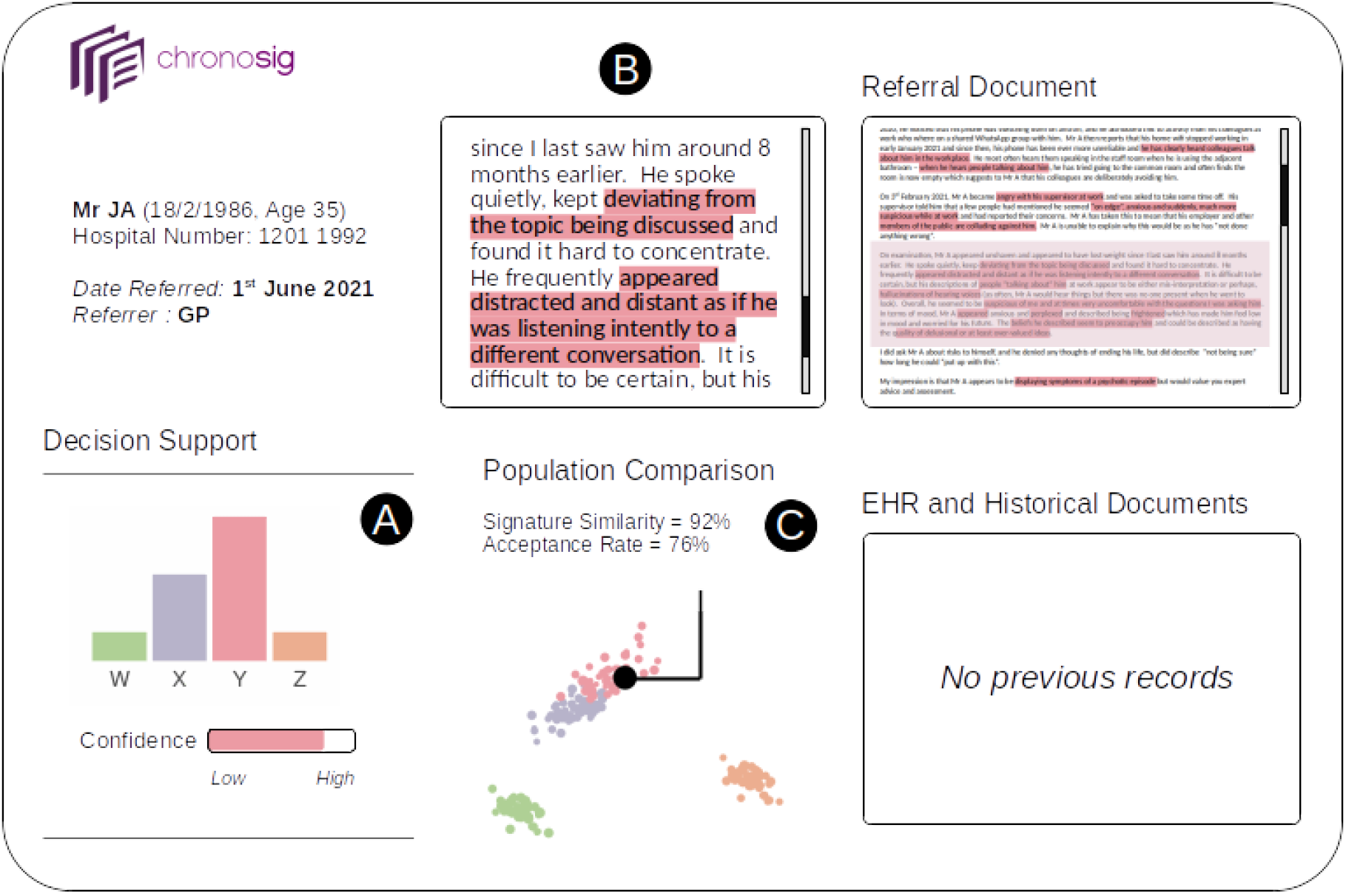
Prototype functionality and user-interface for CHRONOSIG. A patient referral (with no previous psychiatric history) is triaged by the CDS tool which recommends Team Y (with high confidence; panel A). The clinician can use the document map (Referral Document; panel B) to view specific features that support and explain the suggested triage decision. The middle panel (C) visualises the space of all known patient embeddings (with the current patient highlighted) and a numerical estimate of how similar the current patient is to patients previously referred to Team Y alongside an indication of the likelihood this patient would be accepted given their similarity to other patients accepted by Team Y.

**Figure 2:**
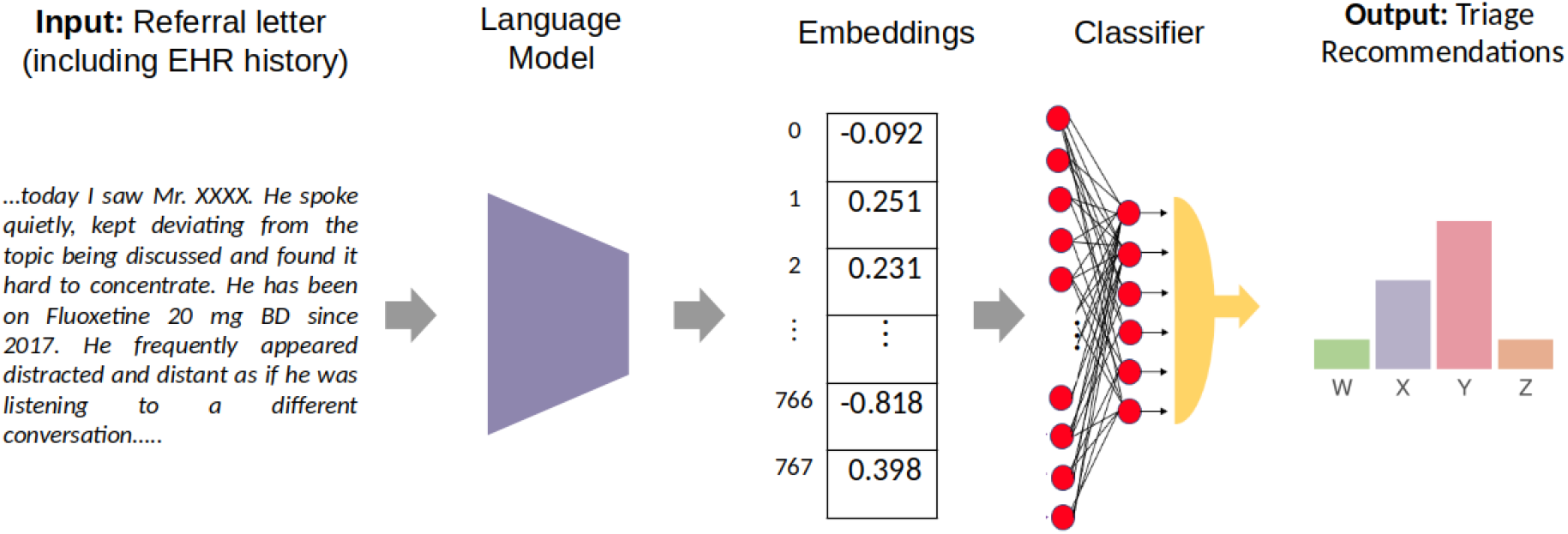
Data Pipeline for CHRONOSIG -- patients’ free-text referral and historical EHR clinical data are used (without *a priori* human annotation) as input to a language model neural network. The resulting numerical representations of patients’ data (denoted “embeddings” or longitudinal fingerprints) are the input to a moderately-sized downstream supervised neural network classifier that learns associations between treatment teams and the embedding representations.

### Patient and Public Co-Production

CHRONOSIG has been developed and funded to provide a patient and public involvement (PPI) work package that runs concurrently with engineering and experimental work. The first PPI deliverable is a stakeholder impact assessment using the FAST (Leslie, 2019) principles (fairness, accountability, sustainability and transparency) in parallel to dataset curation. This output will include PPI (patient and public involvement) definitions of an *acceptable referral* i.e. what characteristics stakeholders expect should be included or excluded from referrals, for example, features of a patient’s presentation which are thought to negatively bias teams in routine practice.

Data curation is the first milestone in CHRONOSIG and the PPI team will direct the technical team to deliver descriptive analyses relevant to the stakeholder impact assessment. For example, requesting analyses of the representation of gender/diagnosis combinations that, historically, have been poorly served by SMH services as well as providing summaries comparing different Trust’s samples to the Office for National Statistics population census data to query representation of minority groups. This output will be made available as meta-data for the benefit of the wider UK-CRIS network.

The representational scheme employed in CHRONOSIG -- namely, longitudinal signatures of EHR text -- aims to provide high fidelity representations of a given patient’s *instance* data (equating to e.g. a referral or an historical segment of their EHR history). A disadvantage of this scheme is that although unlikely, there is a theoretical risk of ‘reverse inference’ where signatures contain enough information to either reconstruct elements of the source free-text or identify the patient indirectly. Using word-vector embeddings (Abdalla et al., 2020) demonstrated that up to 68% of patient name/condition pairs could be recovered (i.e. if the model is built from *identifiable* patient data, associated diagnoses can be gleaned from the representation). For large Transformer language models, again trained on identifiable data, patient name probes generated candidate diagnoses at only the base frequency of each diagnosis in the training data (Lehman et al., 2021) suggesting such models present minimal risk of directly identifying patient/condition pairs. Of course, how NLP models represent broader patient identifiable data (beyond just names) remains an active concern and in CHRONOSIG the PPI group will direct the technical team in stress-testing the signature-generating model to ensure indirect leakage is practically impossible. Additionally, this work will establish that outliers in the space of signatures do not correlate with individual patients or groups of patients with specific protected characteristics (i.e. to prevent undue influence of outliers in the downstream classification model for CDST).

Existing referral and triage processes are not transparent (Chew-Graham et al., 2007), are internal to the SMH team and outcomes and decisions are communicated between the team and the referring party. The National Institute for Health and Care Excellence guidelines on improving patient experiences in mental healthcare (NICE, 2011) recommended studying shared decision making but did not make explicit reference to transparency of the ‘gatekeeping’ role of referral and triage. As a tool for augmented clinical decision making trained on observational data, CHRONOSIG risks recapitulating and entrenching existing practices; given our emphasis on decision making processes we will conduct a patient-led qualitative study during the simulated MDTs. The two primary objectives will be to study existing MDT practices (complementing Chew-Graham *et al*’s work) as well as observing changes in MDT’s practice when the CDST is present. Using semi-structured observation the patient investigator will capture a) what data are being used by the MDT to triage (i.e. patient’s clinical and demographic features) b) whether triage decisions are made transparent and justified with clinical reasoning and c) when there is disagreement among team members, what drives this e.g. patient factors such as diagnostic or risk history alongside MDT factors such as task and process conflict (Jehn & Mannix, 2001).

### Data Curation

The UK-CRIS network (https://crisnetwork.co/uk-cris-programme) enables the curation of datasets of de-identified patient EHR records with well-established operating procedures for secure data curation/storage, ethics, information governance and cyber-security. CHRONOSIG will curate a sample from four demographically different NHS Trusts in the UK-CRIS network for model development and validation with a completely held-out and vaulted sample for model testing.

Patients will be sampled on the basis of types of community teams they were referred to and treated by. The curated dataset will contain unstructured free text records (so-called “progress notes” which capture referral information, details of previous clinical assessments/treatment and administrative contacts with the patient) alongside structured data representing CMHTs the patients were treated by (i.e. a proxy for triage outcomes).

Each patient instance is the unstructured text data from the first time-stamped EHR entry through to the point of a given referral to a secondary care service. A single patient having multiple contacts/treatment episodes with SMH services will be represented by many instances, each one containing the cumulative clinical history from the start of the patient’s EHR record up to the timestamp of a referral or episode of care (reflecting the data a team would have access to when triaging a given referral). When a patient was admitted to hospital (for example, in an emergency) these episodes are not counted as a referral because these are triaged and managed differently from referrals to community services (which make up the majority of secondary mental health care and are the focus of this project). However, the consequent unstructured inpatient clinical data will remain in any future instances if the patient was subsequently referred to community care because inpatient progress data would be available to a triaging MDT.

### Data Bias and Algorithmic Fairness

Similar to data collected by other public sector bodies (ONS, 2018), healthcare records rarely have reliable data recording for ethnicity, gender identity, sexual orientation, relationship status and culture identity. Internationally, people from ethnic groups that are minorities in their domiciled countries are more likely to be admitted for involuntary psychiatric care (Barnett et al., 2019). Surveys conducted with LGBTQ+ communities show susceptibility to specific mental health problems as well as identifying prejudice, stigmatisation and discrimination from healthcare organisations as being reasons people do not seek input for mental health problems (Stonewall, 2018). Together, these factors predict lower representation in our curated dataset and by construction, any statistical model of these properties will be biased. In CHRONOSIG, we will first provide a descriptive audit of available data in UK-CRIS to describe recording for under-represented people compared to population and census data. This will provide meta-data to the wider community making use of UK-CRIS data.

There is no single technical solution to the problem of biased source data. For the derived signature representations, we will use dimensionality reduction methods to visualise networks of signatures with patients explicitly labelled for possessing protected characteristics or being members of under-represented groups -- this enables us to determine if the signature representations can differentially ‘expose’ patients that may be detrimental in the downstream classification (triage outcome) pipeline.

Inferring unobserved and protected characteristic data is demonstrably problematic (Tomasev et al., 2021) -- for example, the risk of unintentional disclosure or an incorrect assumption of predicted gender identity during a triage activity using the CDST. Given we cannot ‘correct’ for under-representation in data sources we will apply principles of *distributive justice* for algorithmic fairness (Rajkomar et al., 2018) specifically:

- we will not defer triage *decisions* to an algorithm -- the CDST delivers transparent *recommendations* whose fairness (and statistical performance) are only evaluated in the context of MDT decision processes (which themselves, may display unfair biases)
- by examining clinical decision making using simulated triage MDTs (described below) we will employ *fairness elicitation* (Jung et al., 2020) as a partial solution to deploying constraints on CDST outputs to promote individually equitable triage for all patients

### Natural Language Processing for Longitudinal Patient Representation

Various NLP methods have been developed and applied to represent and extract clinically meaningful information from chronologically ordered patients’ free-text records (Dalianis, 2018). The common approach was to train an information extraction model (Kormilitzin et al., 2021; Wang et al., 2018) to recognise a predefined set of clinical concepts of interest (e.g. medications, symptoms, health conditions). Once recognised, the concepts were organised as a temporal knowledge graph (Kormilitzin et al., 2020; Leetaru & Schrodt, 2013) that can be queried (Senior et al., 2020).

However, the development of contextual word representations and Transformer-based language models, such as BERT (Devlin et al., 2019) have led to numerous “end-to-end” applications where embedded representations of clinical text inputs are used directly for downstream analytical tasks, such as disease progression modelling and clinical decision support (Bai et al., 2018).

More recently, very large language models (Brown et al., 2020) and task-specific prompting, demonstrated a promising approach to maximise rich information contained in voluminous textual data. However, BERT-like models suffer from the input text length limitation of 512 tokens, whereas the average document length of one year-worth patient’s notes may contain on average 11,000 tokens. Several mitigation strategies were introduced to learn from long texts, including a hierarchical chunking of long texts (Zhang et al., 2020) and sparse attention mechanisms (Zaheer et al., 2020)

We will exploit a recently proposed mathematical methodology of the low-rank tensor approximations (Toth et al., 2020a) to augment the sparse attention mechanism. The new attention mechanism will leverage rich information contained in long chronological EHR to represent patients’ trajectories and ensure the training is computationally efficient. The low-rank tensor approximation for sequential data has already showed promising results (Toth et al., 2020b) on standard benchmark datasets and will allow to capture the long-range dependencies in chronological EHR whereby tokens are separated further apart to learn the longitudinal signature of patients’ health and interventions. The pre-trained language models adapted for long texts, will encode a chronological collection of input free-text patients’ records into a high-dimensional feature vector, representing the longitudinal patients’ signature for downstream predictive tasks.

### Simulated MDTs for Evaluating Augmented Decision Making

Our proposal is that clinical decision making should be *augmented* -- not replaced -- by CDSTs because historical emphasis on automated decision making has under-delivered (Longoni et al., 2019; Nagendran et al., 2020). Clinical decision making requires at the very minimum, knowledge of the risks of triage decisions i.e. a function of the probability of -- and the costs associated with -- each potential outcome. For example, if an MDT rejects a referral, they will be *implicitly* estimating the risks for the patient weighed against those of accepting an inappropriate referral; while one could easily estimate a resource cost of accepting the referral, the cost of catastrophic outcomes -- such as the death of a patient -- cannot be meaningfully captured in the same “currency” (Vickers & Elkin, 2006). Additionally, even in well-calibrated predictive models, assumptions implicit in algorithmic decision rules (i.e. that health resource *cost* is an appropriate proxy for health *needs*) can lead to inequitable outcomes across different racial groups (Obermeyer et al., 2019).

In CHRONOSIG, summary performance metrics (e.g. triage outcome accuracy and error rates) will only be used to evidence that models are learning from the available data. The performance of the tool will be evaluated in controlled experiments where triage decisions from simulated MDTs with and without the CDST are compared. Simulated MDTs will be composed of mental health professionals representative of clinical teams who make triage decisions in secondary mental health care. In a single experiment, patient instances from the vaulted testing sample (i.e. with known triage outcomes but not used during model development) will be presented to both MDTs for triage. The primary outcome will be the between-MDT difference in time spent on each patient’s triage. Secondary outcomes will include analyses of a) within-MDT agreement (homogeneity of the team’s decision making) between-MDT agreement (to establish if triage decisions are automation-biased by having the CDST) and c) identifying pathological triage cases where e.g. the CDST recommendations are consistently at odds with the MDTs decisions. A further qualitative study of the MDT process was described above.

## DISCUSSION

The central tenets of the CHRONOSIG project are:

- Historically, patient and public stakeholders were not involved in process design for referral and triage in SMH care; CDSTs risk recapitulating and entrenching poor practice recorded in the observational EHR data to be used in training algorithms. CHRONOSIG has a mixed-methods work package that oversees and reports on data curation, engineering processes and CDST evaluation from the patient’s perspective.
- Representation of minority and protected groups are known to be biased in observational healthcare data (Rajkomar et al., 2018). CHRONOSIG will explicitly audit source data biases as well as implement methods to dissect and identify biased decisions in the resulting CDSTs.
- Current language model architectures (e.g. ClinicalBERT and Transformers with sparse-attention) are versatile and tractable methods for NLP using text clinical records. CHRONOSIG will attempt to improve on their input sequence length limitation (e.g. 512 - 4096 token strings) to capture long-term dependencies in texts and increase representational capacity for deriving longitudinal signatures.
- Healthcare artificial intelligence (AI) applications have focused on achieving ‘super-human’ performance i.e. the proportion of correct diagnostic classifications made by an AI compared to clinicians. This emphasis led to over-promise, under-delivery and patient mistrust (Longoni et al., 2019; Nagendran et al., 2020). Further, healthcare algorithmic fairness research has demonstrated racial bias (Obermeyer et al., 2019) arising from the choice of decision rule applied to predictive model outputs. Therefore, performance of the CHRONOSIG CDST will be explicitly measured from simulations of triage meetings involving clinicians with and without the tool. CDSTs deliver predictions or recommendations that are inherently uncertain (Joyce & Geddes, 2020) and modern ML methods are mostly opaque “black boxes’’. To verify that a CDST is making clinically-safe decisions (Samek et al., 2017) we require human-interpretable output, including uncertainty estimates, alongside abstractive summaries; in CHRONOSIG, clinicians can interrogate features in the documents that inform triage decisions from the CDST.

### Patient Safety. Governance and Regulatory Factors

For this development project, it would be premature and a risk to patient safety to prospectively evaluate effectiveness with ‘live’ triage cases for clinical use. Instead, validation, testing and initial efficacy data for the CDST will be collected using simulated MDTs with data from historical patients in the UK-CRIS database. The project benefits from the federated UK-CRIS network and inherits the infrastructure and a mature information governance framework. Patient-level EHR data in the UK-CRIS database is pseudonymised and uniquely identifying data (e.g. NHS numbers, dates of birth and names) are masked prior to data being warehoused for research use. Access to UK-CRIS data is strictly controlled to researchers approved by each contributing NHS Trust and there are patient opt-out mechanisms in place. From the perspective of deploying the CDST tool, no source EHR data is required (or stored) for deployment and as described above, risks of reverse inference for patient identifiable data (Abdalla et al., 2020; Lehman et al., 2021) will be investigated thoroughly.

### Deployment at Scale

Almost all modern neural network-based predictive models require substantial computing resources in excess of the typical IT capacity available at a clinician’s desk - a problem well recognised in the NHS (BMA, 2019) but receiving less emphasis in forward-looking state-of-the-art surveys (Joshi & Morley, 2019). Therefore, if this project succeeds and the initial efficacy data is positive, we will migrate the CDST to a cloud-service to facilitate a full parallel-arm prospective controlled trial to include health economic impact. Moving to cloud-based services also mitigates the carbon footprint associated with training language models (Bender et al. 2021) that is essential to the development of responsible AI technology and consistent with the NHS’ ambitions for minimising climate impact.

## Data Availability

Data described in the this paper are only available to authorised and qualified users via the UK-CRIS network

https://crisnetwork.co/

## List of Abbreviations

NHS: National Health Service
EHR: Electronic Health Record
NIHR: National Institute of Health Research
CDST: Clinical Decision Support Tool
UK-CRIS: United Kingdom Clinical Records Interactive Search
NLP: Natural Language Processing
AI: Artificial Intelligence
SMH: Secondary Mental Healthcare
CMHT: Community Mental Health Team
MDT: Multi-Disciplinary Team
ML: Machine Learning
FAST: Fairness, Accountability, Sustainability and Transparency
PPI: Patient and Public Involvement
LGBTQ+: Lesbian, Gay, Bisexual, Transgender, Queer/Questioning and related communities
BERT: Bidirectional Encoder Representations from Transformers
ClinicalBERT: Clinical Bidirectional Encoder Representations from Transformers

## DECLARATIONS

### Ethics approval and consent to participate

This protocol describes research on anonymised patient data in a national database (UK-CRIS) for which the Health Research Authority (in the UK) has issued a waiver in March 2020. Consequently, UK-CRIS projects are not required to have global research ethics committee approval. Individual work-packages in this project are required to be approved by an oversight committee for each request for a subset of data. Therefore, at each milestone work-package in the CHRONOSIG project, requests for specific collections of data will be submitted to each of the four collaborating NHS Trust’s CRIS Oversight Committees to ensure the requested data is an appropriate use of the data under the terms of information governance protocols for UK-CRIS. Participants in the UK-CRIS database are patients receiving secondary mental health care in one of sixteen participating NHS trusts and their anonymised electronic healthcare record data is present in the UK-CRIS database unless they exercise their right to opt-out. Individual participants are not approached for verbal or written consent to participate in this specific study. The UK-CRIS platform has a mature governance framework, including opt-out mechanisms for patients, which can be found at https://crisnetwork.co/governance.

## Consent for publication

Not applicable

## Availability of data and materials

Not applicable

## Competing interests

DWJ provides ad-hoc paid consulting (unrelated to this project) to Akrivia Health, a University of Oxford spinout company that manages the UK-CRIS database. AK is supported in part by GlaxoSmithKline (unrelated to this project).

## Funding

The CHRONOSIG project is funded by the National Institute for Health Research AI for Health and Social Care Programme (Grant Number: AI_AWARD02183)

AC is supported by the National Institute for Health

Research (NIHR) Oxford Cognitive Health Clinical Research Facility, by an NIHR

Research Professorship (grant RP-2017-08-ST2-006) and by the NIHR Oxford Health Biomedical Research Centre (grant BRC-1215-20005).

DWJ is supported by the NIHR Oxford Health Biomedical Research Centre (grant BRC-1215-20005).

The views expressed are those of the authors and not necessarily those of the UK National Health Service, the NIHR, or the UK Department of Health.

## Authors’ contributions

All authors were involved in the design of the study protocol. DWJ, AK and JHH wrote the first draft of the manuscript with all authors contributing to subsequent versions. All authors (DWJ, AK, JHH, AJ, ANH and AC) read and approved the final manuscript.

## Acknowledgements

Not applicable

## Notes

### Funding Statement

This project is funded by the National Institute for Health Research AI for Health and Social Care Programme (Grant Number: AI_AWARD02183)

### Author Declarations

This protocol describes research that will be conducted on anonymised patient data in a national database (UK-CRIS) for which the Health Research Authority (in the UK) has issued a waiver in March 2020.

